# Observational Study of Chlorpromazine in Hospitalized Patients with Covid-19

**DOI:** 10.1101/2020.07.15.20154310

**Authors:** Nicolas Hoertel, Marina Sanchez Rico, Raphaël Vernet, Anne-Sophie Jannot, Antoine Neuraz, Carlos Blanco, Cédric Lemogne, Guillaume Airagnes, Nicolas Paris, Christel Daniel, Alexandre Gramfort, Guillaume Lemaitre, Mélodie Bernaux, Ali Bellamine, Nathanaël Beeker, Frédéric Limosin, On behalf of AP-HP / Universities / INSERM Covid-19 research collaboration and AP-HP Covid CDR Initiative

**Author notes:** Corresponding author Nicolas Hoertel, M.D., M.P.H., Ph.D., Corentin Celton Hospital, AP-HP.Centre, Paris University, 4 parvis Corentin Celton; 92130 Issy-les-Moulineaux, France, /.

## Abstract

On the grounds of its anti-inflammatory and potential antiviral effects, chlorpromazine has been suggested to be effective treatment for Covid-19. We examined the association between chlorpromazine use and respiratory failure among all hospitalized adults with Covid-19 at the 39 Greater Paris University hospitals since the beginning of the epidemic. Study baseline was defined as the date of hospital admission. The primary endpoint was a composite of intubation or death in a time-to-event analysis adjusting for numerous potential confounders. We used a multivariable Cox model with inverse probability weighting according to the propensity score. Of the 12,217 adult inpatients with a positive Covid-19 RT-PCR test included in the analyses, 57 (0.47%) received chlorpromazine. Over a mean follow-up of 20.8 days, the primary endpoint occurred in 29 patients (50.9%) exposed to chlorpromazine and 1,899 patients (15.6%) who were not. In the main analysis, there was a positive significant association between chlorpromazine use and the outcome (HR, 1.67; 95% CI, 1.09 to 2.56, p=0.019), while a Cox regression in a matched analytic sample yielded non-significant association (1.38; 95% CI, 0.91 to 2.09, p=0.123). These findings suggest that chlorpromazine is unlikely to have a clinical efficacy for Covid-19.

## 1. Introduction

Global spread of the novel coronavirus SARS-CoV-2, the causative agent of coronavirus disease 2019 (Covid-19), has created an unprecedented infectious disease crisis worldwide. In the absence of a vaccine or antiviral medications with proven clinical efficacy ^1,2^, the search for an effective treatment for patients with Covid-19 among all available medications is urgently needed ^2,3^.

Chlorpromazine, a dimethylamine derivative of phenothiazine used in the treatment of acute and chronic psychoses ^4^, has been suggested as potential effective treatment for Covid-19 on the grounds of its antiviral and anti-inflammatory effects ^5^. Specifically, several in-vitro studies ^6-8^ showed that chlorpromazine reduces viral replication of coronavirus-229E, MERS-CoV et SARS-CoV-1, possibly through the inhibition of clathrin-mediated endocytosis ^9,10^. Furthermore, several mouse models of sepsis ^11-14^ suggest that this medication is associated with a decrease in pro-inflammatory cytokines, including IL-2, IL4, IFN alpha, TNF, and GM-CSF, and an increase of the anti-inflammatory cytokine IL-10. Short-term use of chlorpromazine is generally well tolerated ^5,15^, although side effects can occur, including QT interval prolongation, extrapyramidal symptoms, dry mouth, dizziness, urine retention, blurred vision, constipation, and hyperprolactinemia ^5,15^.

To our knowledge, no study has examined to date the potential efficacy of chlorpromazine for Covid-19 in clinical populations. Observational studies of patients with Covid-19 taking medications for other indications can help determine their efficacy for Covid-19 and decide which medications should be prioritized for randomized clinical trials, and minimize the risk for patients of being exposed to potentially harmful and ineffective treatments.

To this end, we took advantage of the continuously updated Assistance Publique-Hôpitaux de Paris (AP-HP) Health Data Warehouse, which includes all inpatient visits for Covid-19 to one of the 39 Greater Paris University hospitals since the beginning of the epidemic.

In this report, we examined the association between chlorpromazine use and respiratory failure among adult patients with Covid-19 who have been hospitalized in these medical centers. We hypothesized that chlorpromazine use would be associated with a lower risk of a composite endpoint of intubation or death in a time-to-event analyses that were adjusted for major predictors of respiratory failure and weighted according to propensity scores assessing the probability of chlorpromazine use.

## 2. Methods

### 2.1. Setting

We conducted this study at AP-HP, which comprises 39 hospitals, 23 of which are acute, 20 adult and 3 pediatric hospitals. We included all adults aged 18 years or over who have been admitted to the hospital with Covid-19 from the beginning of the epidemic in France, i.e. January 24^th^, until May 20^th^. In all patients, Covid-19 infection was ascertained by a positive reverse-transcriptase–polymerase-chain-reaction (RT-PCR) test from analysis of nasopharyngeal or oropharyngeal swab specimens. This observational non-interventional retrospective study using routinely collected data received approval from the Institutional Review Board of the AP-HP clinical data warehouse (decision CSE-20-20_COVID19, IRB00011591). AP-HP clinical Data Warehouse initiative ensures patients’ information and consent regarding the different approved studies through a transparency portal in accordance with European Regulation on data protection and authorization n°1980120 from National Commission for Information Technology and Civil Liberties (CNIL). Participants who did not consent to participate in the study were excluded. All procedures related to this work adhered to the ethical standards of the relevant national and institutional committees on human experimentation and with the Helsinki Declaration of 1975, as revised in 2008.

### 2.2. Data sources

We used data from the AP-HP Health Data Warehouse (‘Entrepôt de Données de Santé (EDS)’). This warehouse contains all the clinical data available on all inpatient visits for Covid-19 to any of the 39 AP-HP Greater Paris University hospitals. The data obtained included patients’ demographic characteristics, RT-PCR test results, medication administration data, past and current medication lists, past and current diagnoses, discharge disposition, ventilator use data, and death certificates.

### 2.3. Variables assessed

We obtained the following data for each patient at the time of the hospitalization: sex; age; obesity (defined as having a body-mass index higher than 30 kg/m^2^ or an International Statistical Classification of Diseases and Related Health Problems (ICD-10) diagnosis code for obesity (E66.0, E66.1, E66.2, E66.8, E66.9); self-reported smoking status; any medical condition associated with increased risk of severe SARS-CoV-2 infection ^16-21^, which were coded by practitioners based on ICD-10, including diabetes mellitus (E11), diseases of the circulatory system (I00-I99), diseases of the respiratory system (J00-J99), neoplasms (C00-D49), and diseases of the blood and blood-forming organs and certain disorders involving the immune mechanism (D5-D8); and any medication prescribed according to compassionate use or as part of clinical trials (e.g., hydroxychloroquine, azithromycin, remdesivir, tocilizumab, or sarilumab). To take into account possible confounding by indication bias for chlorpromazine, we recorded whether patients had any current psychiatric disorder, including delirium (F00-F99 and R41.0), and whether they were prescribed any antipsychotic medication other than chlorpromazine, any benzodiazepine or Z-drug, or any other psychotropic medication (i.e., antidepressants or mood stabilizers).

All medical notes and prescriptions are computerized in Greater Paris University hospitals. Medications and their mode of administration (i.e., dosage, frequency, date, condition of intake) were identified from medication administration data or scanned hand-written medical prescriptions, through two deep learning models based on BERT contextual embeddings ^22^, one for the medications and another for their mode of administration. The model was trained on the APmed corpus ^23^, a previously annotated dataset for this task. Extracted medications names were then normalized to the Anatomical Therapeutic Chemical (ATC) terminology using approximate string matching.

### 2.4. Exposure to chlorpromazine

Study baseline was defined as the date of hospital admission. Patients were considered to have been exposed to chlorpromazine if they were under this medication at admission, ascertained by an ongoing medical prescription dated of less than 3 months before hospital admission, or if they received it during the follow-up period before the end of the hospitalization or intubation or death.

### 2.5. Endpoints

The primary endpoint was the time from study baseline to intubation or death. For patients who died after intubation, the primary endpoint was defined as the time of intubation. Patients without an end-point event had their data censored on May 20^th^, 2020.

### 2.6. Statistical analysis

We calculated frequencies and means (± standard deviations (SD)) of each variable described above in patients exposed and not exposed to chlorpromazine and compared them using chi-square tests or Welch’s t-tests.

To examine the association of chlorpromazine use with the primary composite endpoint (i.e., intubation or death), we performed Cox proportional-hazards regression models. Weighted Cox regression models were used when the proportional hazards assumption was not met. To help account for the nonrandomized prescription of chlorpromazine and reduce the effects of confounding, the primary analysis used propensity score analysis with inverse probability weighting ^24,25^. The individual propensities for chlorpromazine prescription were estimated by a multivariable logistic regression model that included sex, age, obesity, smoking status, any medical condition, any medication prescribed according to compassionate use or as part of a clinical trial, any current psychiatric disorder (including delirium), any antipsychotic medication other than chlorpromazine, any benzodiazepine or Z-drug, and any other psychotropic medication. In the inverse-probability-weighted analysis, the predicted probabilities from the propensity-score model were used to calculate the stabilized inverse-probability-weighting weight ^24^. Association between chlorpromazine use and the primary endpoint was then estimated using a multivariable Cox regression model using the inverse-probability-weighting weights. Kaplan-Meier curves were performed using the inverse-probability-weighting weights ^26^, and their pointwise 95% confidence intervals were estimated using the nonparametric bootstrap method ^27^.

We conducted sensitivity analyses, including a multivariable Cox regression model comprising as covariates the same variables as the inverse-probability-weighted analysis, and a univariate Cox regression model in a matched analytic sample. For this latter analysis, we selected five controls for each exposed case, based on the same variables used for both the inverse-probability-weighted analysis and the multivariable Cox regression. To reduce the effects of confounding, optimal matching was used in order to obtain the smallest average absolute distance across all these characteristics between each exposed patient and its five corresponding non-exposed matched controls.

Finally, within the group of patients exposed to chlorpromazine, we tested the association of daily dosage and duration of exposure (dichotomized into ‘prescription that began during the hospitalization’ and ‘prescription that started before hospitalization’) with the primary endpoint.

For all significant associations, we performed residual analyses to assess the fit of the data, check assumptions, including the proportional hazards assumption, and examined the potential influence of outliers. To improve the quality of result reporting, we followed the recommendations of The Strengthening the Reporting of Observational Studies in Epidemiology (STROBE) Initiative.^28^ Statistical significance was fixed a priori at p<0.05. All analyses were conducted between June 5^th^ and June 15^th^ in R software version 2.4.3.

## 3. Results

### 3.1. Characteristics of the cohort

Of the 16,170 hospitalized adult patients with a positive Covid-19 RT-PCR test, a total of 3,953 patients (24.4%) were excluded because of missing data. Of the remaining 12,217 adult inpatients, 57 patients (0.47%) were exposed to chlorpromazine, at a mean dosage of 79.5 mg per day (SD=71.6; median=50 mg; range: 6.0 mg to 300.0 mg), and this exposure started after hospital admission in most of them (84.2%, n=48) (**Figure 1**).

**Figure 1.**
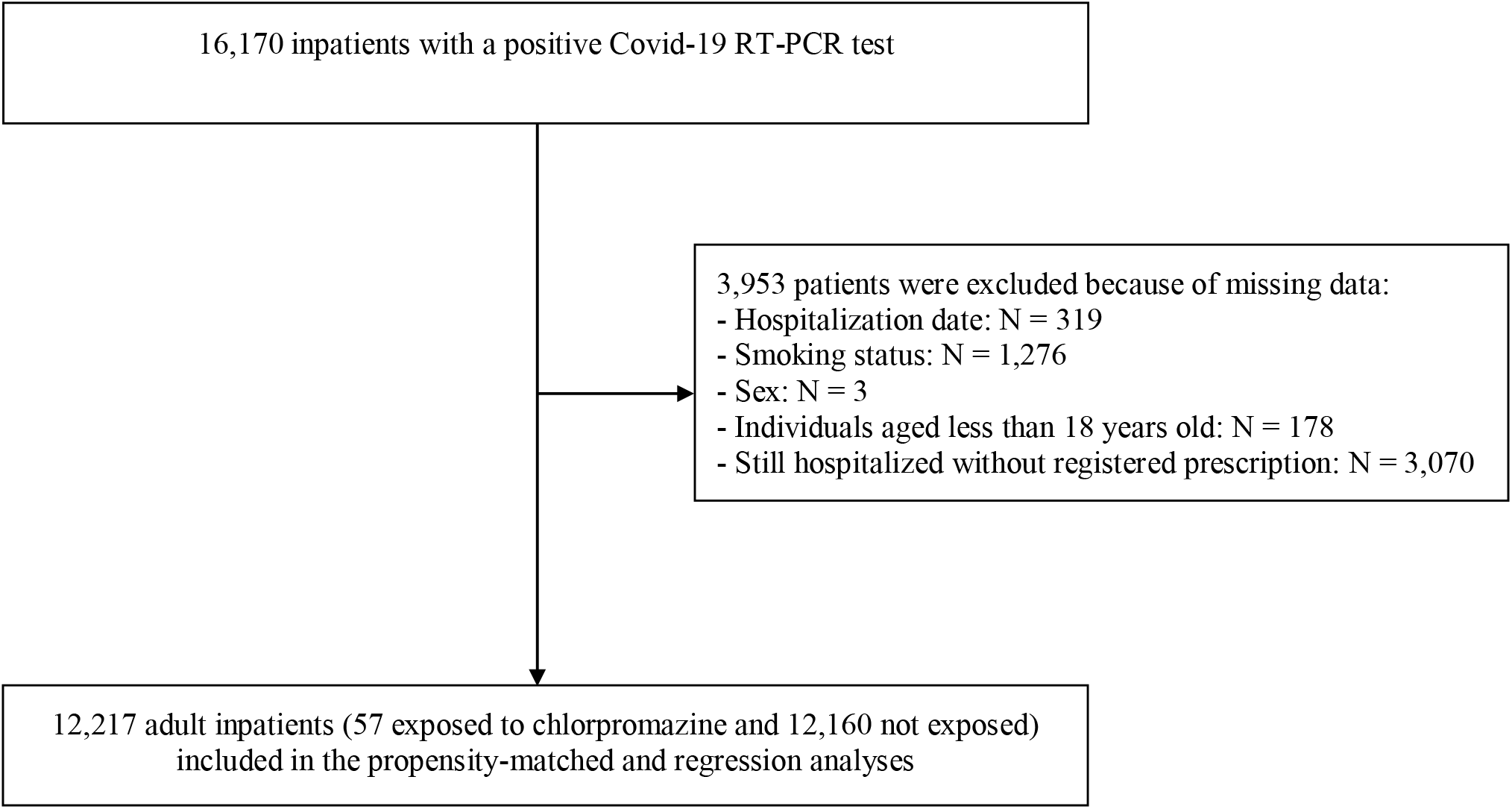
Study cohort.

Over a mean follow-up of 17.8 days (SD=24.0; median=5 days; range: 1 day to 117 days), 1,624 patients (13.3%) had a primary end-point event prior to the completion of data collection on May 20^th^. In patients exposed to chlorpromazine, the mean follow-up was 21.1 days (SD=21.7; median=12 days; range: 1 day to 83 days), while it was of 18.7 days (SD=24.3; median=6 days; range: 1 day to 117 days) for those who were not exposed to chlorpromazine.

The distribution of the patients’ characteristics by chlorpromazine exposure status is shown in **Table 1**. In the full sample, chlorpromazine exposure significantly differed according to presence of obesity, any medical condition, any current psychiatric disorder, any psychotropic medication other than chlorpromazine, any benzodiazepine or Z-drug, and any other psychotropic medication, and the direction of associations indicated greater medical severity of people receiving chlorpromazine than those who did not. After applying the propensity score weights, these differences were substantially reduced and remained significant only for any current psychiatric disorder, any benzodiazepine or Z-drug, and any other psychotropic medication (**Table 1**).

**Table 1.**
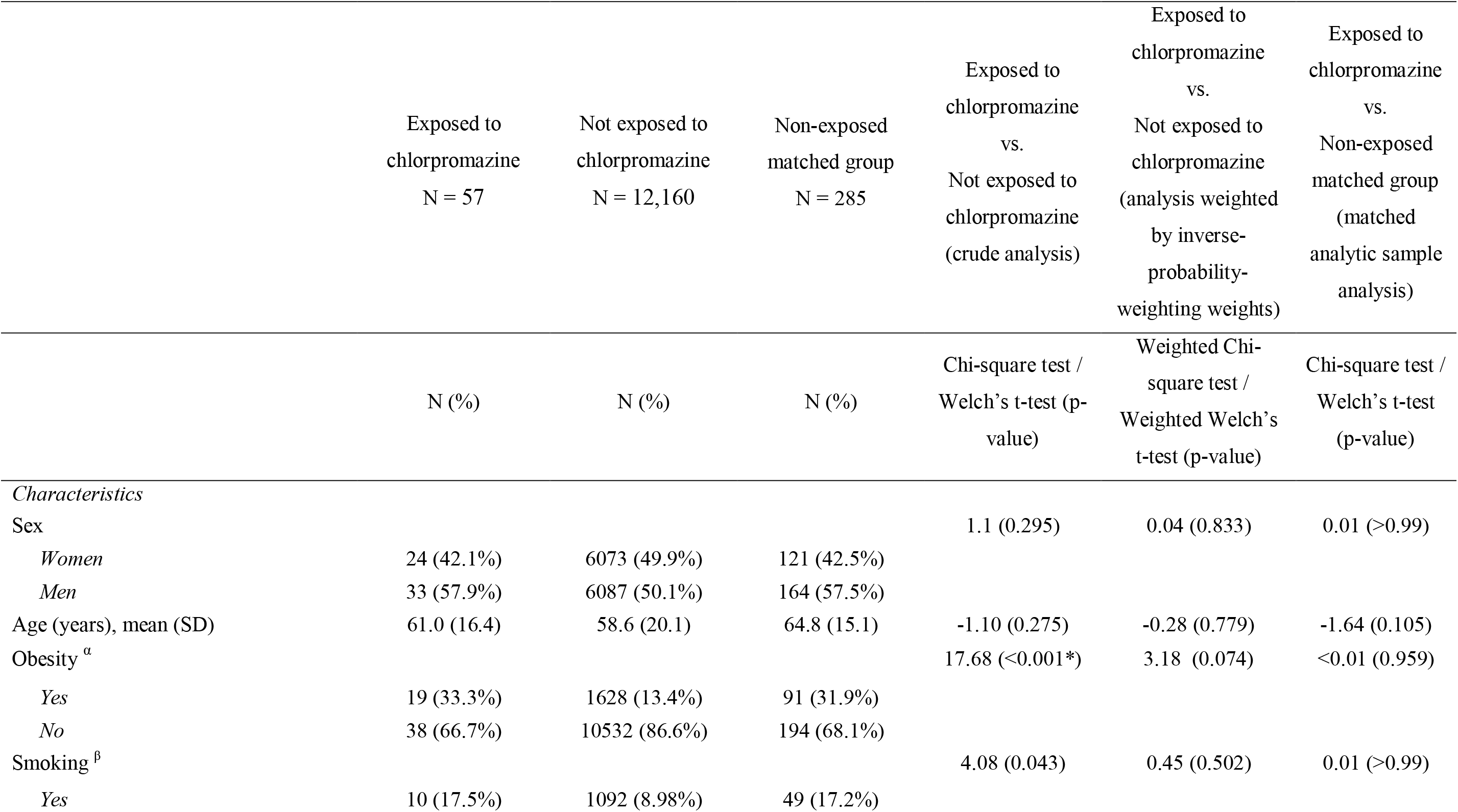
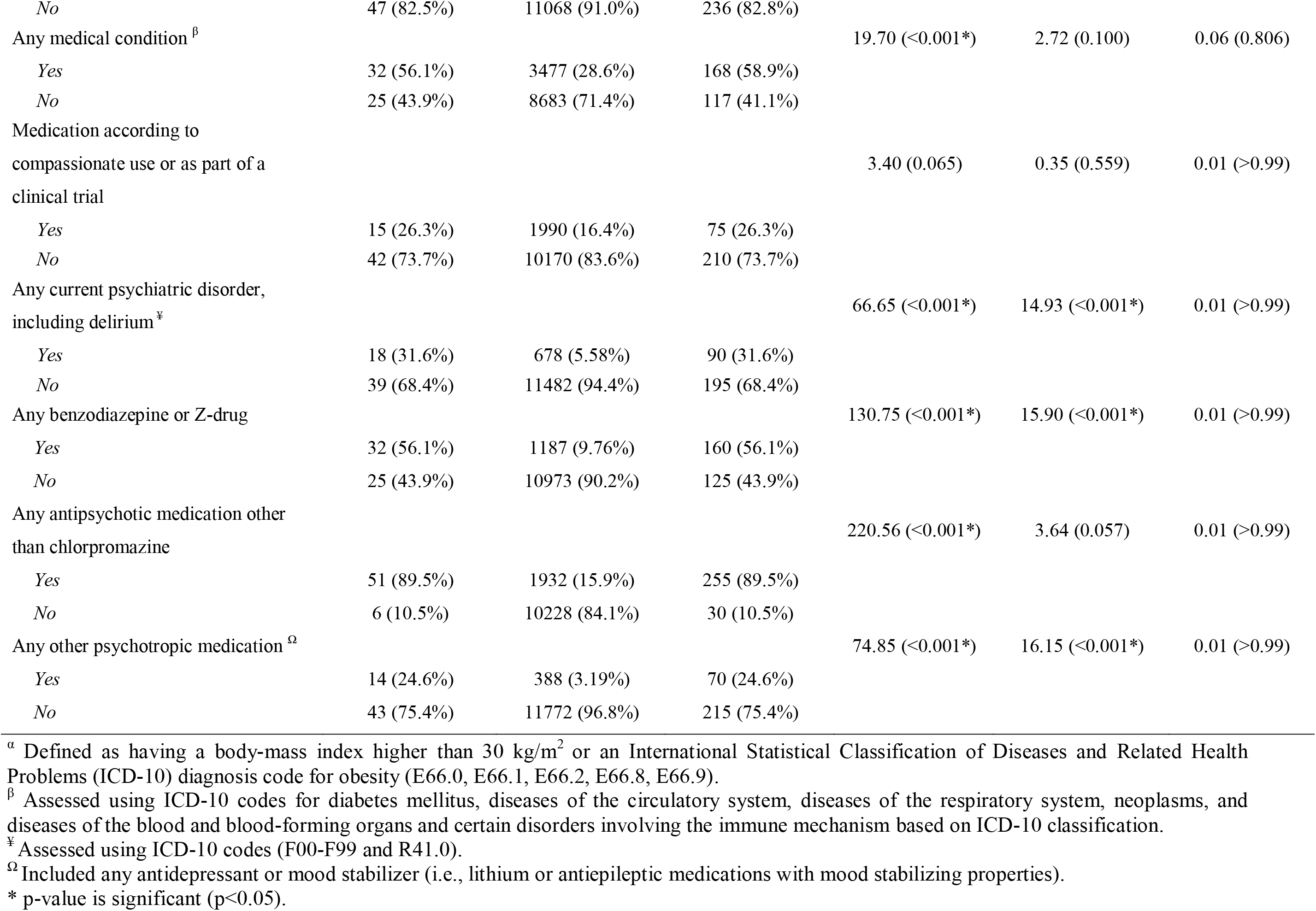
Characteristics of patients receiving or not receiving chlorpromazine in the matched and unmatched analytic samples.

|  | Exposed to<br>chlorpromazine<br>N = 57 | Not exposed to<br>chlorpromazine<br>N = 12,160 | Non-exposed<br>matched group<br>N = 285 | Exposed to<br>chlorpromazine<br>vs.<br>Not exposed to<br>chlorpromazine<br>(crude analysis) | Exposed to<br>chlorpromazine<br>vs.<br>Not exposed to<br>chlorpromazine<br>(analysis weighted<br>by inverse-probability-weighting weights) | Exposed to<br>chlorpromazine<br>vs.<br>Non-exposed<br>matched group<br>(matched<br>analytic sample<br>analysis) |
| --- | --- | --- | --- | --- | --- | --- |
|  | N (%) | N (%) | N (%) | Chi-square test /<br>Welch's t-test (p-value) | Weighted Chi-square test /<br>Weighted Welch's<br>t-test (p-value) | Chi-square test /<br>Welch's t-test<br>(p-value) |
| *Characteristics* |  |  |  |  |  |  |
| Sex |  |  |  | 1.1 (0.295) | 0.04 (0.833) | 0.01 (>0.99) |
| *Women* | 24 (42.1%) | 6073 (49.9%) | 121 (42.5%) |  |  |  |
| *Men* | 33 (57.9%) | 6087 (50.1%) | 164 (57.5%) |  |  |  |
| Age (years), mean (SD) | 61.0 (16.4) | 58.6 (20.1) | 64.8 (15.1) | -1.10 (0.275) | -0.28 (0.779) | -1.64 (0.105) |
| Obesity <sup>a</sup> |  |  |  | 17.68 (<0.001*) | 3.18 (0.074) | <0.01 (0.959) |
| *Yes* | 19 (33.3%) | 1628 (13.4%) | 91 (31.9%) |  |  |  |
| *No* | 38 (66.7%) | 10532 (86.6%) | 194 (68.1%) |  |  |  |
| Smoking <sup>β</sup> |  |  |  | 4.08 (0.043) | 0.45 (0.502) | 0.01 (>0.99) |
| *Yes* | 10 (17.5%) | 1092 (8.98%) | 49 (17.2%) |  |  |  |
| *No* | 47 (82.5%) | 11068 (91.0%) | 236 (82.8%) |  |  |  |
| Any medical condition <sup>β</sup> |  |  |  | 19.70 (<0.001*) | 2.72 (0.100) | 0.06 (0.806) |
| *Yes* | 32 (56.1%) | 3477 (28.6%) | 168 (58.9%) |  |  |  |
| *No* | 25 (43.9%) | 8683 (71.4%) | 117 (41.1%) |  |  |  |
| Medication according to<br>compassionate use or as part of a<br>clinical trial |  |  |  | 3.40 (0.065) | 0.35 (0.559) | 0.01 (>0.99) |
| *Yes* | 15 (26.3%) | 1990 (16.4%) | 75 (26.3%) |  |  |  |
| *No* | 42 (73.7%) | 10170 (83.6%) | 210 (73.7%) |  |  |  |
| Any current psychiatric disorder,<br>including delirium <sup>¥</sup> |  |  |  | 66.65 (<0.001*) | 14.93 (<0.001*) | 0.01 (>0.99) |
| *Yes* | 18 (31.6%) | 678 (5.58%) | 90 (31.6%) |  |  |  |
| *No* | 39 (68.4%) | 11482 (94.4%) | 195 (68.4%) |  |  |  |
| Any benzodiazepine or Z-drug |  |  |  | 130.75 (<0.001*) | 15.90 (<0.001*) | 0.01 (>0.99) |
| *Yes* | 32 (56.1%) | 1187 (9.76%) | 160 (56.1%) |  |  |  |
| *No* | 25 (43.9%) | 10973 (90.2%) | 125 (43.9%) |  |  |  |
| Any antipsychotic medication other<br>than chlorpromazine |  |  |  | 220.56 (<0.001*) | 3.64 (0.057) | 0.01 (>0.99) |
| *Yes* | 51 (89.5%) | 1932 (15.9%) | 255 (89.5%) |  |  |  |
| *No* | 6 (10.5%) | 10228 (84.1%) | 30 (10.5%) |  |  |  |
| Any other psychotropic medication <sup>Ω</sup> |  |  |  | 74.85 (<0.001*) | 16.15 (<0.001*) | 0.01 (>0.99) |
| *Yes* | 14 (24.6%) | 388 (3.19%) | 70 (24.6%) |  |  |  |
| *No* | 43 (75.4%) | 11772 (96.8%) | 215 (75.4%) |  |  |  |

<sup>α</sup> Defined as having a body-mass index higher than 30 kg/m<sup>2</sup> or an International Statistical Classification of Diseases and Related Health Problems (ICD-10) diagnosis code for obesity (E66.0, E66.1, E66.2, E66.8, E66.9).
<sup>β</sup> Assessed using ICD-10 codes for diabetes mellitus, diseases of the circulatory system, diseases of the respiratory system, neoplasms, and diseases of the blood and blood-forming organs and certain disorders involving the immune mechanism based on ICD-10 classification.
<sup>¥</sup> Assessed using ICD-10 codes (F00-F99 and R41.0).
<sup>Ω</sup> Included any antidepressant or mood stabilizer (i.e., lithium or antiepileptic medications with mood stabilizing properties).
\* p-value is significant (p<0.05).

In the matched analytic sample comprising 342 patients (i.e., 57 patients exposed to chlorpromazine and 285 patients from the matched group), there were no significant differences in any characteristic according to chlorpromazine exposure (**Table 1**).

### 3.2. Study endpoint

Respiratory failure occurred in 23 patients (40.4%) who received chlorpromazine and 1,619 patients (13.3%) who did not (**Table 2**). There was a significant positive association between chlorpromazine use and the composite primary endpoint in both the crude, unadjusted analysis (hazard ratio (HR), 2.85; 95% CI, 1.97 to 4.12, p<0.001) and the primary multivariable analysis with inverse probability weighting (HR, 1.67; 95% CI, 1.09 to 2.56, p=0.019) (**Figure 2**; **Table 2**). In sensitivity analyses, the multivariable Cox regression model in the full sample yielded a similar result (hazard ratio (HR), 1.96; 95% CI, 1.34 to 2.86, p<0.001). However, this association was not significant in the univariate Cox regression model in the matched analytic sample (hazard ratio (HR), 1.38; 95% CI, 0.91 to 2.09, p=0.123), for which differences across all characteristics between exposed and non-exposed cases were not significant (**Figure 2**; **Table 2**).

**Table 2.** Associations between chlorpromazine use and the endpoint of intubation or death in the full sample and in the matched analytic sample.

| Analysis | Intubation or death |
| --- | --- |
| ***Full sample*** |  |
| Number of events / Number of patients (%) | 1,328 / 12,217 (10.9%) |
| Chlorpromazine | 29 / 57 (50.9%) |
| No chlorpromazine | 1,899 / 12,160 (15.6%) |
| Crude analysis – hazard ratio (95% CI; p-value) | 2.85 (1.97 – 4.12; <0.001*) |
| Multivariable analysis – hazard ratio (95% CI; p-value) | 1.96 (1.34 – 2.86, <0.001*) |
| Propensity score analysis with inverse probability weighting – hazard ratio (95% CI; p-value) | 1.67 (1.09 – 2.56; 0.019*) |
| ***Matched analytic sample*** |  |
| Number of events / Number of patients (%) | 130 / 342 (38.0%) |
| Chlorpromazine | 29 / 57 (50.9%) |
| No chlorpromazine | 101 / 285 (35.4%) |
| Crude analysis – hazard ratio (95% CI; p-value) | 1.38 (0.91 – 2.09; 0.123) |

\* p-value is significant (p<0.05).

**Figure 2.**
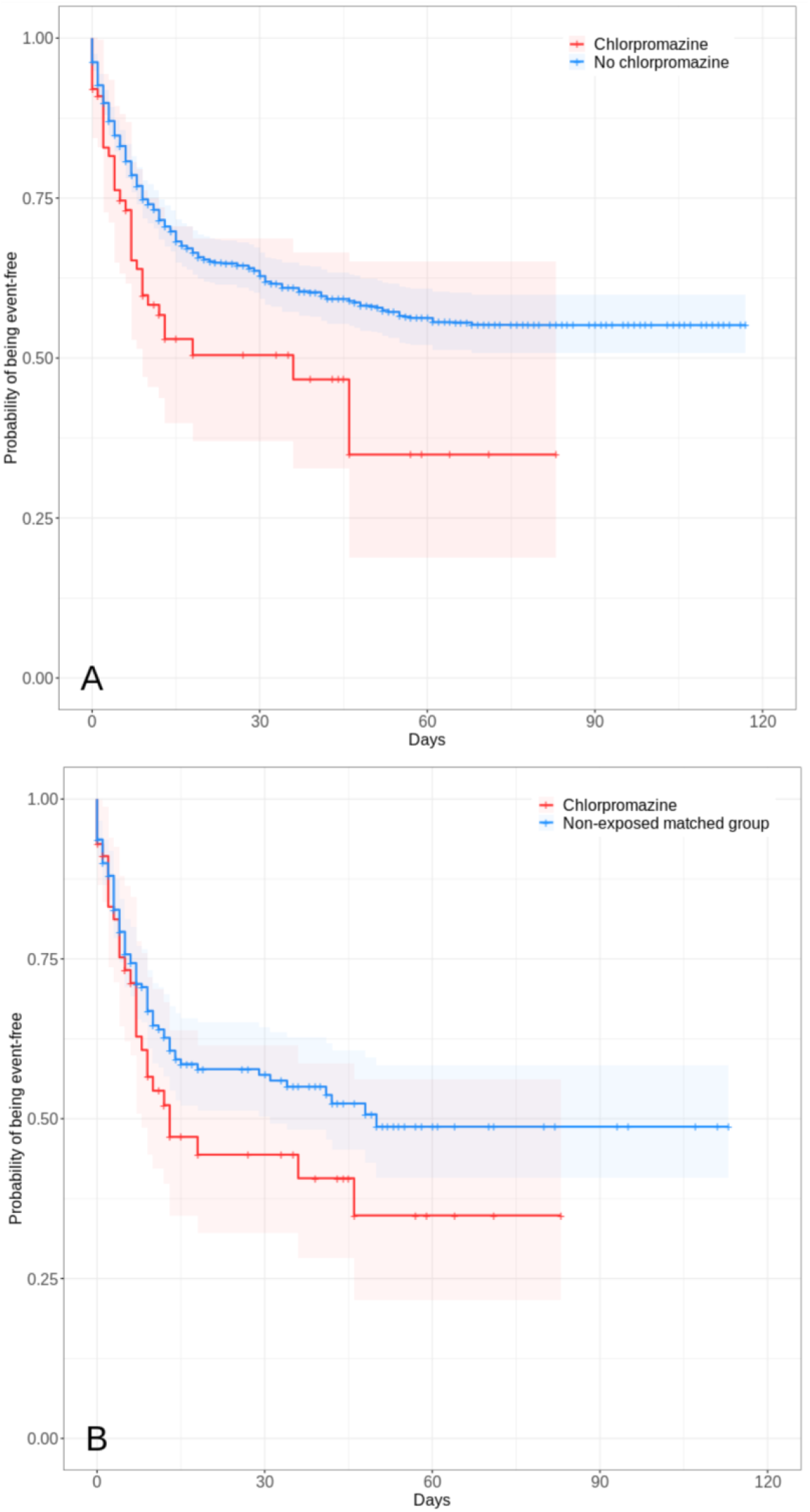
Freedom from the composite endpoint of intubation or death in the full sample (N=12,217) (A) and in the matched analytic sample (N=342) (B) of hospitalized patients with Covid-19 according to chlorpromazine exposure. ^¥^. *Note:* The shaded areas represent pointwise 95% confidence intervals. ^¥^ Patients were considered to be exposed to chlorpromazine if they were under this medication at admission, ascertained by an ongoing medical prescription dated of less than 3 months before hospital admission, or if they received it during the follow-up period before the end of the hospitalization or intubation or death (N=57).

When restricting the analyses to the 48 patients who received chlorpromazine only during the hospitalization, the association between chlorpromazine use and the primary endpoint was significant in the propensity score weighted analysis (HR, 2.00; 95% CI, 1.31 to 3.02, p=0.001), but not in the matched analytic sample analysis (HR, 1.53; 95% CI, 1.00 to 2.37, p=0.051) (**Supplemental eFigure 1**). When restricting the analyses to the 9 patients exposed to chlorpromazine in the 3 months prior to the hospitalization only, this association was not significant in the propensity score weighted analysis (HR, 0.86; 95% CI, 0.25 to 2.90, p=0.808) or in the matched analytic sample analysis (HR, 0.74; 95% CI, 0.23 to 2.32, p=0.602) (**Supplemental eFigure 2**)

A post-hoc analysis indicated that in the full sample, we had 80% power to detect hazard ratios for chlorpromazine treatment of at least 2.38/0.42 for the primary endpoint.

Finally, there were no significant associations of daily chlorpromazine dosage or duration of exposure with the primary endpoint [HR (95% CI), 0.99 (0.98-1.01), p=0.191; and HR (95% CI), 2.73 (0.64-11.75), p=0.177, respectively].

## Discussion

In this observational multicenter study involving a large number of inpatients with Covid-19, exposure to chlorpromazine was not significantly associated with risk of intubation or death. Although these findings should be interpreted with caution due to the observational design, the wide confidence intervals for estimates, and the fact that this is, to our knowledge, the first study examining the association of chlorpromazine use with respiratory failure in a clinical population of patients with Covid-19, our results suggest that chlorpromazine is unlikely to have a clinical efficacy for Covid-19.

In the analyses, we tried to minimize the effects of confounding in several different ways. First, we used a multivariable regression model with inverse probability weighting to minimize the effects of confounding by indication ^24,25^. We also performed sensitivity analyses, including a multivariable Cox regression model and a univariate Cox regression model in a matched analytic sample, which respectively showed a significant positive association between use of chlorpromazine and the primary endpoint and a non-significant association, giving strength to the conclusion that chlorpromazine in unlikely to reduce risk of intubation or death.

Second, although some amount of unmeasured confounding may remain, our analyses adjusted for numerous potential confounders, including sex, age, obesity, smoking status, any medical condition, any medication prescribed according to compassionate use or as part of a clinical trial, any current psychiatric disorder (including delirium), any prescribed antipsychotic medication other than chlorpromazine, any benzodiazepine or Z-drug, and any other psychotropic medication. Finally, the lack of significant associations of daily chlorpromazine dosage or duration of exposure with the endpoint of intubation or death further supports our conclusion.

Additional limitations of our study include missing data for some variables and potential for inaccuracies in the electronic health records, such as the possible lack of documentation of illnesses or medications, or the misidentification of treatment mode of administration (e.g., dosage, frequency), especially for hand-written medical prescriptions. However, results remained unchanged after using multiple imputation to account for missing data (available on request). Furthermore, patients under chlorpromazine were prescribed a relatively low dosage, i.e., 75 mg per day (SD=4.3), and its antiviral properties might be observable at higher dosages. However, we did not find a significant association between dosage and the primary endpoint. In addition, despite the multicenter design, our results may not be generalizable to other settings or regions. Finally, it is possible that chlorpromazine prescription was motivated mainly by the presence of agitation, which might be a marker of severity of Covid-19. However, the analyses adjusted for current psychiatric disorders, including delirium, and non-psychiatric medical conditions.

In this observational study involving patients with Covid-19 who had been admitted to the hospital, chlorpromazine use was not associated with a lower risk of intubation or death. The results suggest that chlorpromazine is unlikely to have a clinical efficacy for Covid-19.

## Data Availability

Data from the AP-HP Health Data Warehouse can be obtained at https://eds.aphp.fr//.

## Acknowledgments

The authors warmly thank the EDS APHP Covid consortium integrating the APHP Health Data Warehouse team as well as all the APHP staff and volunteers who contributed to the implementation of the EDS-Covid database and operating solutions for this database.

Collaborators EDS APHP Covid consortium: Pierre-Yves ANCEL, Alain BAUCHET, Nathanaël BEEKER, Vincent BENOIT, Mélodie BERNAUX, Ali BELLAMINE, Romain BEY, Aurélie BOURMAUD, Stéphane BREANT, Anita BURGUN, Fabrice CARRAT, Charlotte CAUCHETEUX, Julien CHAMP, Sylvie CORMONT, Christel DANIEL, Julien DUBIEL, Catherine DUCLOAS, Loic ESTEVE, Marie FRANK, Nicolas GARCELON, Alexandre GRAMFORT, Nicolas GRIFFON, Olivier GRISEL, Martin GUILBAUD, Claire HASSEN-KHODJA, François HEMERY, Martin HILKA, Anne Sophie JANNOT, Jerome LAMBERT, Richard LAYESE, Judith LEBLANC, Léo LEBOUTER, Guillaume LEMAITRE, Damien LEPROVOST, Ivan LERNER, Kankoe LEVI SALLAH, Aurélien MAIRE, Marie-France MAMZER, Patricia MARTEL, Arthur MENSCH, Thomas MOREAU, Antoine NEURAZ, Nina ORLOVA, Nicolas PARIS, Bastien RANCE, Hélène RAVERA, Antoine ROZES, Elisa SALAMANCA, Arnaud SANDRIN, Patricia SERRE, Xavier TANNIER, Jean-Marc TRELUYER, Damien VAN GYSEL, Gaël VAROQUAUX, Jill Jen VIE, Maxime WACK, Perceval WAJSBURT, Demian WASSERMANN, Eric ZAPLETAL.

## Authorship

NH designed the study, performed statistical analyses, and wrote the first draft of the manuscript. MSR performed statistical analyses and critically revised the manuscript. RV contributed to statistical analyses and critically revised the manuscript for scientific content. FL, NB and ASJ contributed to study design and critically revised the manuscript for scientific content. NB, ASJ, AN, NP, CD, AG, GL, MB, and AB contributed to database build process. AN, CB, CL, GA, NP, CD, AG, GL, MB, and AB critically revised the manuscript for scientific content.

## Data Availability Statement

Data from the AP-HP Health Data Warehouse can be obtained at https://eds.aphp.fr//.

## Disclaimer

The views and opinions expressed in this report are those of the authors and should not be construed to represent the views of any of the sponsoring organizations, agencies, or the US government.

